# Validation of Immunoscore for Prognostic Stratification in HPV-associated Oropharyngeal Cancer: An International Multicenter Study

**DOI:** 10.64898/2026.04.08.26350238

**Authors:** D.H. Nguyen, A. Majdi, F. Marliot, V. Houtart, A. Kirilovsky, A. Hijazi, T. Fredriksen, N. de Sousa Carvalho, A.-S. Bach, A.-L. Gaultier, E. Fabiano, S. Kreps, E. Tartour, H. Péré, D. Veyer, P. Blanchard, H. K. Angell, F. Pagès, H. Mirghani, J. Galon

## Abstract

**Background:** Treatment optimization in HPV-associated oropharyngeal cancer (OPSCC) remains challenging, as recent de-escalation trials have shown limited success. Current patient selection strategies based on smoking history and TNM classification are insufficient, highlighting the need for robust, standardized prognostic biomarkers. We report the first validation of the Immunoscore (IS) for prognostic stratification in HPV-associated OPSCC.

**Patients and methods:** We analyzed 191 HPV-associated (p16+ and HPV DNA/RNA+) OPSCC patients from an international multicenter cohort (2015–2024), comprising a French monocentric retrospective training cohort (N = 48) and three validation cohorts: French monocentric retrospective (N = 48), French multicenter prospective (N = 50), and US multicenter retrospective (N = 45). IS is a standardized digital pathology assay quantifying CD3lJ and CD8lJ densities in tumor cores and invasive margins, with cut-offs defined in the training cohort and validated across cohorts. Associations with disease-free survival (DFS), time to recurrence (TTR) and overall survival (OS) were assessed, alongside 3’RNA-seq and sequential immunofluorescence profiling of immune composition.

**Results:** Median age 65; 80% male; 74% smokers; 66% T1-2; 82% N0-1 (AJCC8^th^). IS-High patients demonstrated superior 3-year DFS in the training and validation cohorts 1-3 (all log-rank *P* < 0.05). Multivariable analysis identified IS-Low as the strongest independent risk factor for DFS (HR 9.03; 95% CI: 4.02-20.31; *P* < 0.001). The model combining IS with clinical factors showed higher predictive accuracy for DFS (C-index 0.82) than clinical variables alone (0.7; *P* < 0.0001). Similar findings were observed for TTR and OS. IS-High tumors showed markedly higher enrichment of lymphoid and myeloid immune cell populations, contrasting with immune-poor signatures in IS-Low tumors.

**Conclusions:** IS is a robust biomarker that outperforms standard clinical variables in both prognostic and predictive accuracy. The enriched cytotoxic immune infiltrate in IS-High tumors explains favorable outcomes and supports their suitability for treatment de-escalation. Prospective validation is warranted.

## INTRODUCTION

For the past decades, the incidence of human papillomavirus (HPV)–associated oropharyngeal cancer (OPSCC) has risen sharply, now comprising 70–80% of new cases in North America and Northern Europe.^1–3^ Despite being a biologically distinct disease with superior outcomes to those of HPV-negative OPSCCs, the standard of care remains uniform, involving high-dose chemoradiotherapy (CRT) or surgery plus adjuvant therapy.^4–7^ While curative, these regimens are associated with substantial toxicity, including mucositis, xerostomia, osteoradionecrosis and long-term dysphagia, driving an urgent need for safe de-escalation strategies.^4,5^

However, recent efforts toward therapeutic de-escalation have encountered significant setbacks. Phase II–III randomized trials that replaced cisplatin with cetuximab, investigated reduced-intensity CRT or combined nivolumab with lower-dose radiotherapy (RT) have resulted in compromised overall survival (OS) and locoregional control, failed to demonstrate non-inferiority or yielded inconclusive results.^6–12^ These challenges underscore that HPV-associated OPSCC is not a clinically homogeneous entity, as approximately 15 to 25% of patients experience disease progression within 2 to 3 years despite standard treatment.^13,14^ Current selection criteria for de-escalation, relying predominantly on the TNM 8^th^ edition classification and smoking history, are inadequate, as they fail to capture the intrinsic biological heterogeneity of the HPV-associated OPSCC population.^15^

Risk stratification may be improved by leveraging features of the tumor microenvironment (TME), particularly tumor-infiltrating lymphocytes (TILs), which are critical determinants of clinical outcomes across solid tumors.^16,17^ Although the prognostic value of TILs in HPV-associated OPSCC has been documented,^18,19^ the lack of a standardized scoring system has hindered clinical application. The Immunoscore (IS) is a standardized clinical digital pathology assay that quantifies CD3^+^ and CD8^+^ T-cells in the tumor core (CT) and invasive margin (IM). To our knowledge, it is the first and only internationally validated assay for quantifying lymphocyte infiltration.^20,21^ In colorectal cancer, IS has been validated as a prognostic tool that outperforms the AJCC/UICC TNM staging system.^21^ Accordingly, immune assessment has been integrated into the 2019 WHO classification,^22^ and IS testing is recommended by both the European Society for Medical Oncology (ESMO) and the Pan-Asian adapted ESMO Clinical Practice Guidelines.^23,24^

In this study, we report the first international multicenter validation of the consensus IS in HPV-associated OPSCC. Transcriptomic profiling and sequential immunofluorescence (seqIF) were used to further characterize the TME and immune populations associated with IS. We hypothesized that IS could stratify patients into distinct risk groups, potentially identifying candidates for therapeutic de-escalation.

## PATIENTS AND METHODS

### Study Design and Patient Cohorts

We conducted a multicenter, international, retrospective-prospective cohort study involving 191 HPV-associated OPSCC patients treated between 2015 and 2024 (**Figure 1A**). The study consisted of a training phase followed by a three-step validation phase. The training cohort consisted of 48 consecutive patients retrospectively treated at European Hospital Georges Pompidou (HEGP) (Paris, France) from 2017 to 2019, used exclusively to establish IS groups. The validation phase utilized three independent datasets: Validation cohort 1 (V1), comprising 48 consecutive patients retrospectively treated at HEGP (2020-2022); Validation cohort 2 (V2), a prospective multicenter cohort of 50 patients recruited from eight centers in France treated from 2021-2024 and Validation cohort 3 (V3), a retrospective cohort of 45 patients treated in the US between 2015 and 2018. The conduct and reporting adhered to the REMARK guidelines for prognostic biomarker studies (**Supplementary Table S1)**.^25^

**Figure 1.**
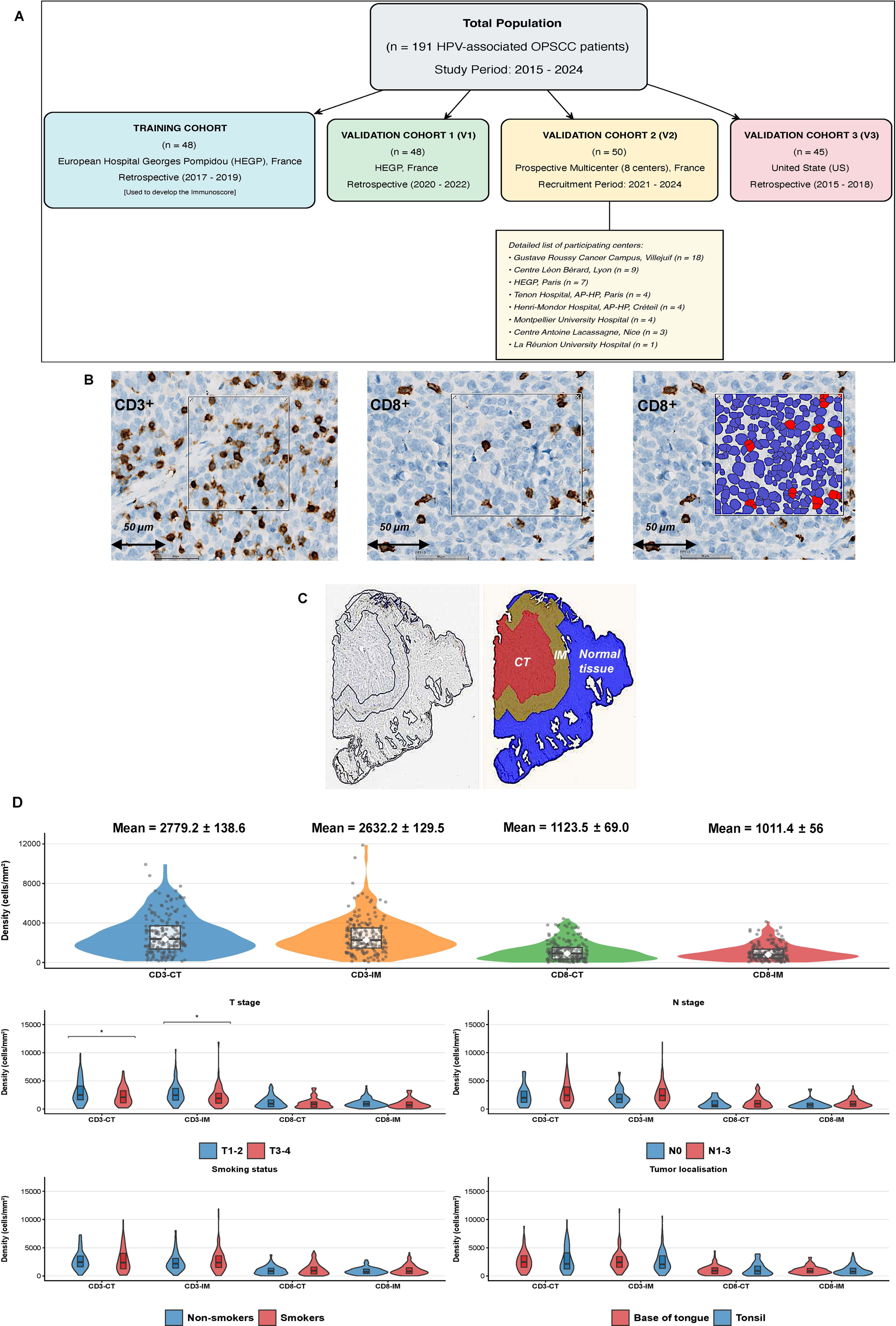
Study design, patient cohorts, and quantitative assessment of CD3□/CD8□ T-cell infiltration (A) Study design and patient cohorts. (B) Representative images were analyzed using HALO™ software, with CD3□/CD8□ cells shown in brown. Automated detection of CD8L□ cells (red) was performed using a predefined algorithm. (C) Tissue segmentation illustrating the tumor core (CT, red), invasive margin (IM, brown) and adjacent normal tissue (blue). (D) Densities of CD3□ and CD8□ cells in the CT and IM regions and their correlation with clinical variables. * *P* < 0.05; ** *P* < 0.01; *** *P* < 0.001; **** *P* < 0.0001.

### Ethical Considerations

This study was approved by the Ethics Committee of the AP-HP Center (CERAPHP; IRB registration number: #00011928). Written informed consent was obtained from all patients for the use of their biological samples and clinical data. For cohort V3, tissue samples were collected with written informed consent and supplied by the AstraZeneca Biobank (UK), approved by the Human Tissue Authority (License No. 12109) and the National Research Ethics Service (REC No. 17/NW/0207).

### Inclusion and Exclusion Criteria

Eligible adults (≥18 years) had histologically confirmed squamous cell carcinoma of the oropharynx (tonsil or base of tongue) and received curative-intent treatment (surgery ± adjuvant therapy or definitive RT/CRT), according to current standard treatment guidelines.^26,27^ Pre-treatment formalin-fixed paraffin-embedded (FFPE) tumor tissues from biopsy or resection specimens were used for analysis. Exclusion criteria included distant metastasis at diagnosis (M1), prior head and neck malignancy, contraindications to standard CRT due to poor performance status, prior treatment with immune checkpoint inhibitors (ICIs) or inadequate tissue samples for analysis.

### Definition of HPV Status

Only samples satisfying both of the following conditions were considered HPV-driven: positive p16INK4a immunohistochemistry (IHC), defined as >70% strong nuclear and cytoplasmic staining (CINtec Histology, Roche MTM laboratories);^28^ and positive HPV DNA or RNA, confirmed by PCR (Allplex II HPV28 Detection; Seegene, Republic of Korea) or HPV mRNA *in situ* hybridization (RNAscope 2.5 HD, Advanced Cell Diagnostics, USA).

### Sample Preparation

FFPE tissue blocks containing both the CT and the IM were selected by experienced pathologist at our center. Ten consecutive 4-µm sections were cut from each block: one for hematoxylin and eosin (H&E) staining, three for IS assay and seqIF and the remaining for RNA extraction.

### Immunoscore protocol

IS was performed according to our standardized protocol as previously described ^21^ in the coordinating center (Immunomonitoring Platform, HEGP, Paris, France). IHC was carried out on an automated platform (BenchMark XT, Ventana) using anti-CD3 (2GV6) and anti-CD8 (C8/144B) antibodies (**Supplementary File S2)**, with DAB detection, hematoxylin counterstaining and slide digitization at ×20 (NanoZoomer-XR scanner).

Cut-offs for CD3L and CD8L cell densities in the CT and IM were set at the 25^th^ percentile of their respective distributions in the training cohort, consistent with the validated consensus IS definition of the immune-low category in colorectal cancer.^21^ For the scoring algorithm, each variable (CD3-CT, CD3-IM, CD8-CT, CD8-IM) was assigned a binary score (1 if above the cut-off, 0 if below), and the final IS (0–4) was calculated as their sum. Patients were subsequently stratified according to two classification schemes:

**-** Two-category Immunoscore: IS-Low (0-2) vs. IS-High (3-4).
**-** Three-category Immunoscore: IS-Low (0-1), IS-Intermediate (2-3) and IS-High (4).

All cut-offs and category definitions for IS were fixed prior to further analysis in the training and validation cohorts.

### SeqIF protocol

SeqIF was performed on 56 patients (16 from training cohort, 19 from V1 and 21 from V2), including 34 IS-High (60.7%) and 22 IS-Low (39.3%) cases. FFPE 4-µm sections underwent heat-induced epitope retrieval (pH 9, 102 °C, 60 min) and were processed for seqIF on the automated COMET™ platform (Lunaphore Technologies, Switzerland), enabling iterative multiplex immunofluorescence on a single tissue section.^29^ The antibody panel included: CD4L T cells, Granzyme B (GZMB)L cytotoxic cells, CD56L natural killer (NK) cells, FOXP3L regulatory T cells, CD20L B cells, MUM1L plasma cells, CD11cL dendritic cells, CD68L macrophages, myeloperoxidase (MPO)L granulocytes, pan-cytokeratin (CK)L epithelial cells and α-SMAL (myo)fibroblasts (**Figure 3D**). Detailed information on the antibody used is provided in **Supplementary File S2**.

### Digital Image Analysis

Images were analyzed using HALO™ software v4.1.5 (Indica Labs, USA) by operators blinded to clinical data. Predefined CD3 and CD8 thresholds for IS were applied across all samples, with cell densities (cells/mm²) were quantified separately in the CT and IM (**Figure 1B-C**). For seqIF, thresholds were automatically set by the software based on immunofluorescence intensity and cell densities were calculated exclusively within pathologist-annotated tumor regions (**Supplementary Figure S3**).

### Transcriptomic Profiling

Pathologist annotations on H&E slides guided the macrodissection of tumoral tissue from FFPE sections, which was then used for RNA extraction with the NucleoSpin total RNA FFPE kit (Macherey-Nagel, Germany). For the training, V1 and V2 cohorts, libraries were prepared using the QuantSeq 3’ mRNA-Seq Kit (Lexogen, Vienna, Austria) and sequenced on an Illumina NextSeq 500 (10 million reads/sample). Cell-type enrichment analysis was performed using the xCell 2.0 package in R-Studio (v4.2.0).^30^ Gene Ontology (GO) and pathway enrichment analyses were conducted using ShinyGO 0.85 to identify biological processes enriched among differentially expressed genes.^31^ FDR-adjusted *P* < 0.05 was considered significant. For the cohort V3, gene expression was assessed using the NanoString nCounter PanCancer Immune Profiling Panel (NanoString Technologies, US). Differential gene expression analysis was performed using DESeq2. Genes with FDR-adjusted *P* < 0.05 and |logL fold change| ≥ 0.5 were considered significant.

### Statistical Analysis

The primary endpoint was the prognostic value of IS for 3-year DFS, with secondary endpoints including 3-year time to recurrence (TTR) and OS. DFS events were defined as local, regional, distant recurrence or death from any cause; TTR events included only cancer recurrences and OS events were deaths from any cause. Patients without events were censored at last follow-up. Survival was estimated using the Kaplan-Meier method and compared with the log-rank test. Univariate and multivariable Cox proportional hazards models assessed IS as an independent prognostic factor, adjusting for smoking status, cT/cN stage (AJCC 8^th^ edition), and treatment modality. The multivariable model was applied either to the entire cohort (*N* = 191) or restricted to the validation cohorts only (*N* = 143). The predictive accuracy of the models was evaluated using Harrell’s Concordance Index (C-index) and the integrated Area Under the Curve (iAUC) with 1,000 bootstrap resamples for internal validation. Risk prediction models were compared using the likelihood ratio *P*-value. Model quality was assessed using the Akaike Information Criterion (AIC), with ΔAIC >10 indicating a substantial improvement between models. A two-sided *P*-value < 0.05 was considered statistically significant. All analyses were performed using R-Studio (v4.2.0).

## RESULTS

### Cohort Characteristics

The detailed characteristics of the 191 patients are summarized in **Table 1**. The mean age was 64.7 years (SD 11.0), and 80.1% of patients were male. Current or former smokers accounted for 74.3% of the cohort; pack-year (PY) data were available for 185 of 191 patients (96.9%), with 61% exceeding 10 PY. Primary tumor sites were distributed between the tonsil (54.5%) and the base of tongue (45.5%). 65.9% had T1-2 disease and 81.7% were N0-1. Treatments included surgery with adjuvant RT/CRT (64.4%), definitive RT/CRT (26.7%) or surgery alone (8.9%). The median follow-up time was 37.6 months (IQR, 26.4-59.7). Across cohorts, significant differences were observed in age, smoking exposure, tumor site, T stage and follow-up duration (**Table 1**).

**Table 1.**
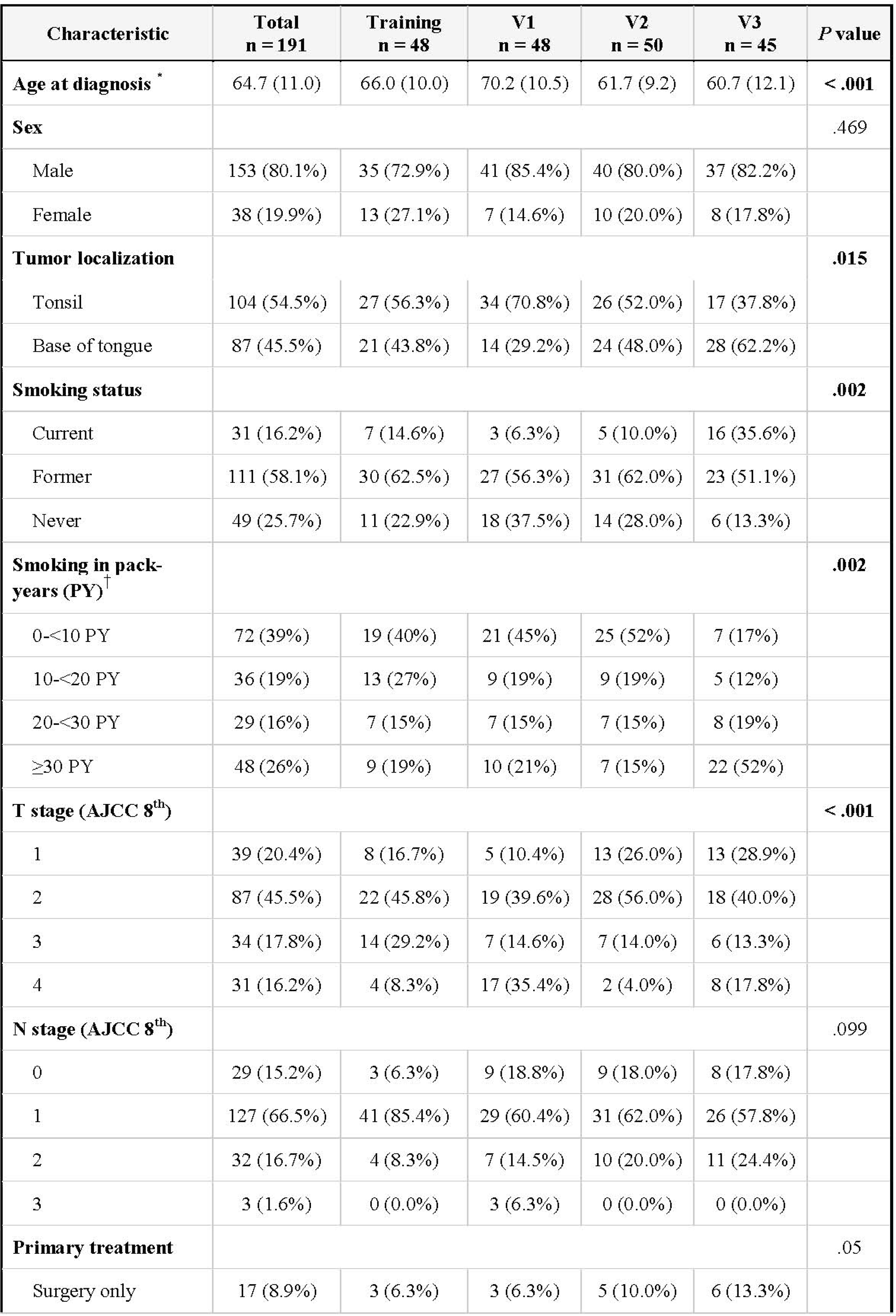
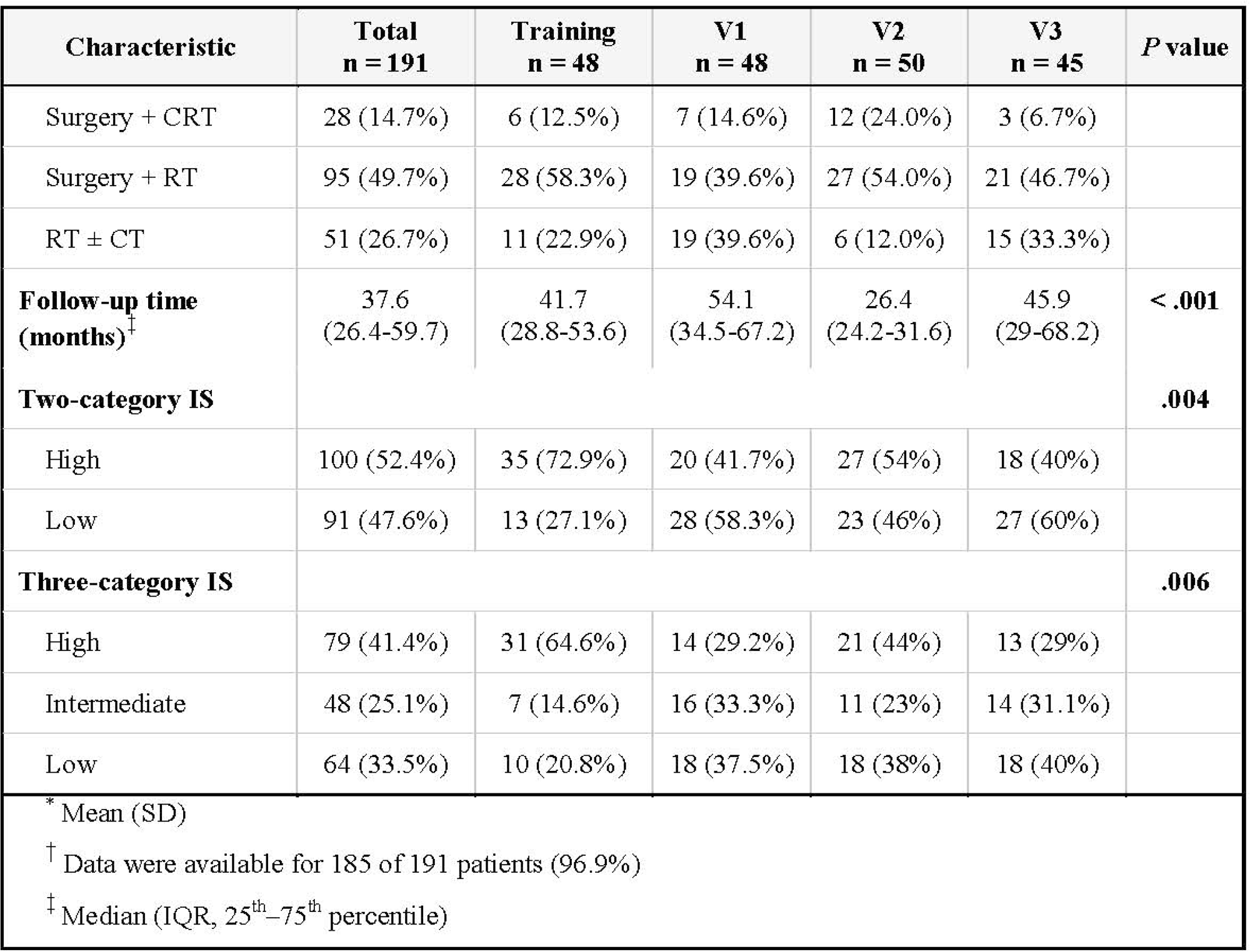
Demographic and clinical characteristics of the study population.

### Lymphocyte Densities and Clinical Correlations

Mean densities and distributions of CD3L and CD8L cells are shown in **Figure 1D**. The densities of CD3L cells in both the CT and the IM were significantly higher in T1-2 tumors than in T3-4 tumors (*P* = 0.037 and *P* = 0.016, respectively). No significant association was observed with smoking status, tumor localization, or N stage **(Figure 1D).**

### Distribution and Prognostic Impact of IS

Cut-off values for each immune cell density were set at the 25^th^ percentile of the respective distributions in the training cohort. Applying these predefined cut-offs to the total population, 52.4% were IS-High (*N* = 100) and 47.6% IS-Low (*N* = 91). The prevalence of the IS-High status differed significantly across cohorts: 72.9% in the training cohort, 41.7% in V1, 54% in V2 and 40% in V3 (*P* = 0.004) **(Table 1)**. No significant differences in clinical characteristics were observed between IS-High and IS-Low patients (**Supplementary Table S4).**

In the overall cohort, IS-Low patients had significantly worse 3-year DFS than IS-High patients (univariate Cox HR 8.57; 95% CI 3.87-18.96; *P* < 0.001) (**Figure 2A-B**; **Supplementary File S5)**. The 3-year DFS was 93.4% (95% CI: 87.8-99.3) for IS-High compared to 58.9% (95% CI: 49.3-70.3) for IS-Low (*P* < 0.0001) (**Figure 2B**). This finding was consistent across all cohorts, with IS-High patients showing significantly better 3-year DFS in the training (*P* = 0.002), V1 (*P* = 0.003), V2 (*P* = 0.012) and V3 (*P* = 0.008) cohorts (**Figure 2A**). Similar patterns were observed for TTR and OS. IS-High patients demonstrated superior outcomes compared to IS-Low patients, including 3-year TTR (93.4% [95% CI: 87.8-99.3] vs. 62.4% [95% CI: 53-73.5]) and 3-year OS (97.3% [95% CI: 93.5-100] vs. 78.1% [95% CI: 69.5-87.8]) (all *P* < 0.0001) (**Supplementary Figure S6).** Importantly, IS effectively stratified DFS, TTR, and OS in both clinically low-risk (T1–3, N0–2, non-active smokers; *N* = 136) and high-risk patients (*N* = 55) (all *P* < 0.01) **(Supplementary Figure S7)**. Survival did not differ significantly between IS-High patients with low versus high clinical risk (all *P* > 0.05). Across treatment subgroups, IS effectively stratified survival in patients receiving definitive RT ± CT (*N* = 51) and in those undergoing surgery with adjuvant RT ± CT (*N* = 123; all *P* < 0.001). No significant differences were observed in the surgery-only group (*N* = 17), likely due to the small sample size (**Supplementary Figure S8**). Using the three-category IS, the observed 3-year DFS rates were 91.6% (95% CI: 84.5-99.2), 73.5% (95% CI: 61.6-87.9) and 61.1% (95% CI: 49.9-74.7) for the High, Intermediate and Low groups, respectively (*P* < 0.0001) (**Figure 2C**). Consistent findings were observed for TTR (*P* < 0.0001) and OS (*P* = 0.001) (**Supplementary Figure S6).**

**Figure 2.**
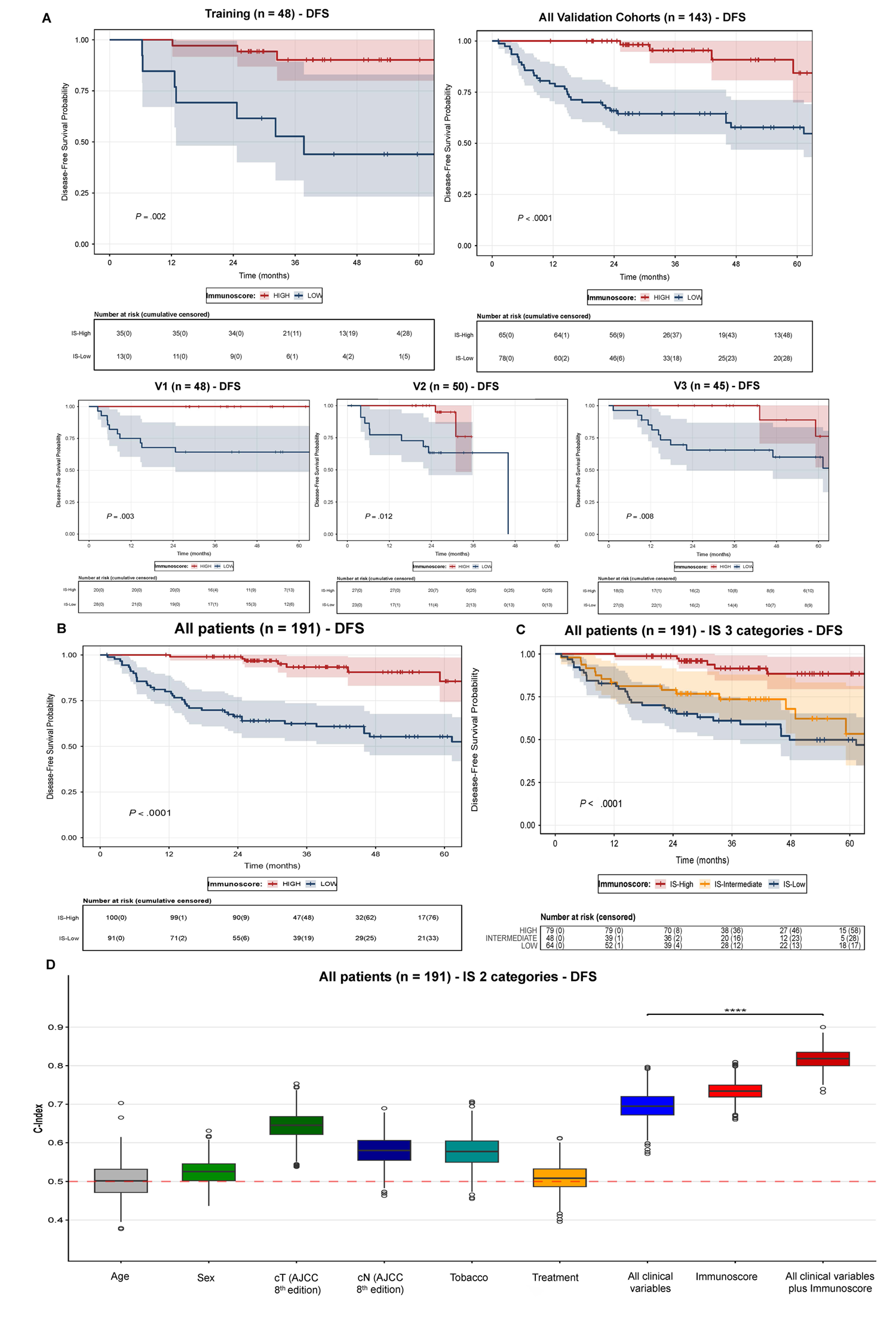
Prognostic value of IS for DFS. (A) Kaplan-Meier curves for DFS according to IS (High vs Low) in the training cohort and each validation cohort. (B) Kaplan–Meier curves for DFS according to IS (High vs Low) in the overall population. (C) Kaplan–Meier curves for DFS according to the three-category IS classification (High, Intermediate, Low). (D) Box plots show the distribution of Harrell’s Concordance Index (C-index) for 3-year DFS across different prognostic models, estimated using 1,000 bootstrap resamples. The log-likelihood ratio test comparing the clinical model alone with the clinical model plus IS is shown. * *P* < 0.05; ** *P* < 0.01; *** *P* < 0.001; **** *P* < 0.0001.

### Multivariable Analysis and Predictive Accuracy of IS

In the overall cohort, multivariable Cox regression adjusted for smoking status, cT/cN stage (AJCC 8^th^) and treatment modalities identified IS-Low status as the strongest independent prognostic factor for DFS (HR 9.03; 95% CI: 4.02-20.31; *P* < 0.001) **(Table 2)**. This effect exceeded that of active smoking (HR 3.12; 95% CI: 1.34-7.25; *P* = 0.008), cT3-4 stage (HR 2.32; 95% CI: 1.29-4.14; *P* = 0.005), cN stage, and treatment modality. Cumulative tobacco exposure (PY) was not prognostic for DFS. IS was also the strongest independent prognostic factor for TTR (HR 7.45; 95% CI: 3.27-16.97; *P* < 0.001) and OS (HR 6.77; 95% CI: 2.57-17.87; *P* < 0.001). The prognostic value of IS was confirmed in multivariable analyses performed in the validation cohorts only (*N* = 143) (all *P* < 0.001 for DFS, TTR, and OS).

**Table 2.**
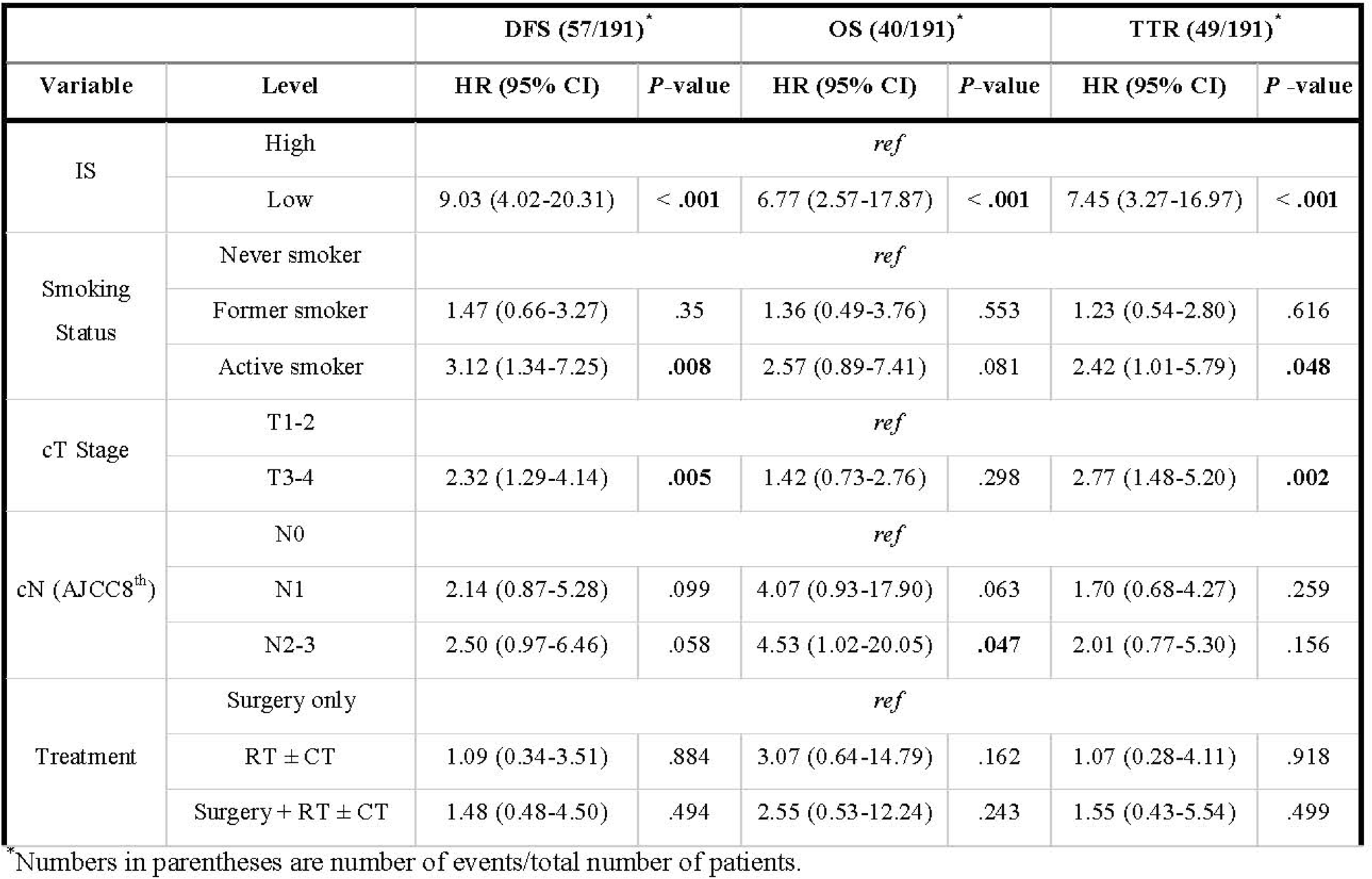
Multivariable Cox proportional hazards regression analyses for DFS, OS and TTR adjusted for clinical covariates.

In terms of predictive accuracy, the standard clinical model incorporating all clinical variables yielded C-index values of 0.7, 0.71 and 0.71 for DFS, TTR and OS, respectively (**Figure 2D**; **Supplementary File S5)**. The IS model alone achieved higher C-index (0.73, 0.73 and 0.71). The integrated model (IS + all clinical variables) yielded the highest accuracy, with C-index of 0.82, 0.82 and 0.8 for DFS, TTR and OS, significantly outperforming the clinical model (log-likelihood ratio test *P* < 0.0001). For 3-year DFS, iAUC values were 0.73, 0.75, and 0.84 for the clinical, IS-only, and combined models, respectively. ΔAIC between the clinical-only and combined models was 41, 32.4, and 16.5 for DFS, TTR, and OS, demonstrating that adding IS substantially improved model performance (**Supplementary File S5**).

### Transcriptomic and seqIF analysis

Across the training, V1, and V2 cohorts (*N* = 146), IS-High tumors (*N* = 82) showed a rich cytotoxic transcriptomic profile, enriched for T cells & TCR signaling (*CD2/5/7, CD3D/E/G, CD8A/B, TRAC/TRDC/TRBC1/2/TRGC2, LCK, CD40LG*), NK/NK_T (*GNLY, GZMA/B, PRF1, KLRG1/B/F1/D1, NKG7, IFNG*), immune checkpoints (*TBX21, ICOS, FOXP3, CTLA4*), plasma/B cells (*CD19, CD27/37/53, BANK1, BLK, MS4A1, PAX5, TNFRSF17, FCRL2*) and myeloid infiltration (*CLEC4E, SIGLEC1, TFEC, CD1B, CD86, CCR7*) (all adjusted *P* < 0.05) (**Figure 3A**; **Supplementary File S10)**. In contrast, IS-Low tumors (*N* = 64) showed a more fibrotic and pro-angiogenic signatures, with upregulated stromal components (*COL1A1/2, COL3A1, COL4A1/6, FN1, LOX/LOXL2, SPARC, TGFBI, TIMP1*) and angiogenesis genes (*VEGFA/C*) (all adjusted *P* < 0.05) (**Figure 3A**). Cell-type enrichment confirmed higher adaptive immune populations in IS-High tumors (CD8L, CD4L T cells, B cells, dendritic cells, macrophages) versus enrichment of epithelial, keratinocyte and mesenchymal stromal cells in IS-Low tumors (all adjusted *P* < 0.05) (**Figure 3B**). Additionally, GO and pathway analyses showed that IS-High tumors were enriched for immune-related processes (antigen receptor signaling, lymphocyte differentiation, adaptive immune response), whereas IS-Low tumors were associated with extracellular matrix organization, vasculature development, cell migration, and adhesion (all FDR-adjusted *P* < 0.05) (**Figure 3C)**. These patterns were reproduced in the independent V3 cohort (**Supplementary Figure S9, Supplementary File S10**), consolidating the “immune-hot” versus “immune-cold” transcriptomic phenotypes of IS-High and IS-Low tumors.

**Figure 3.**
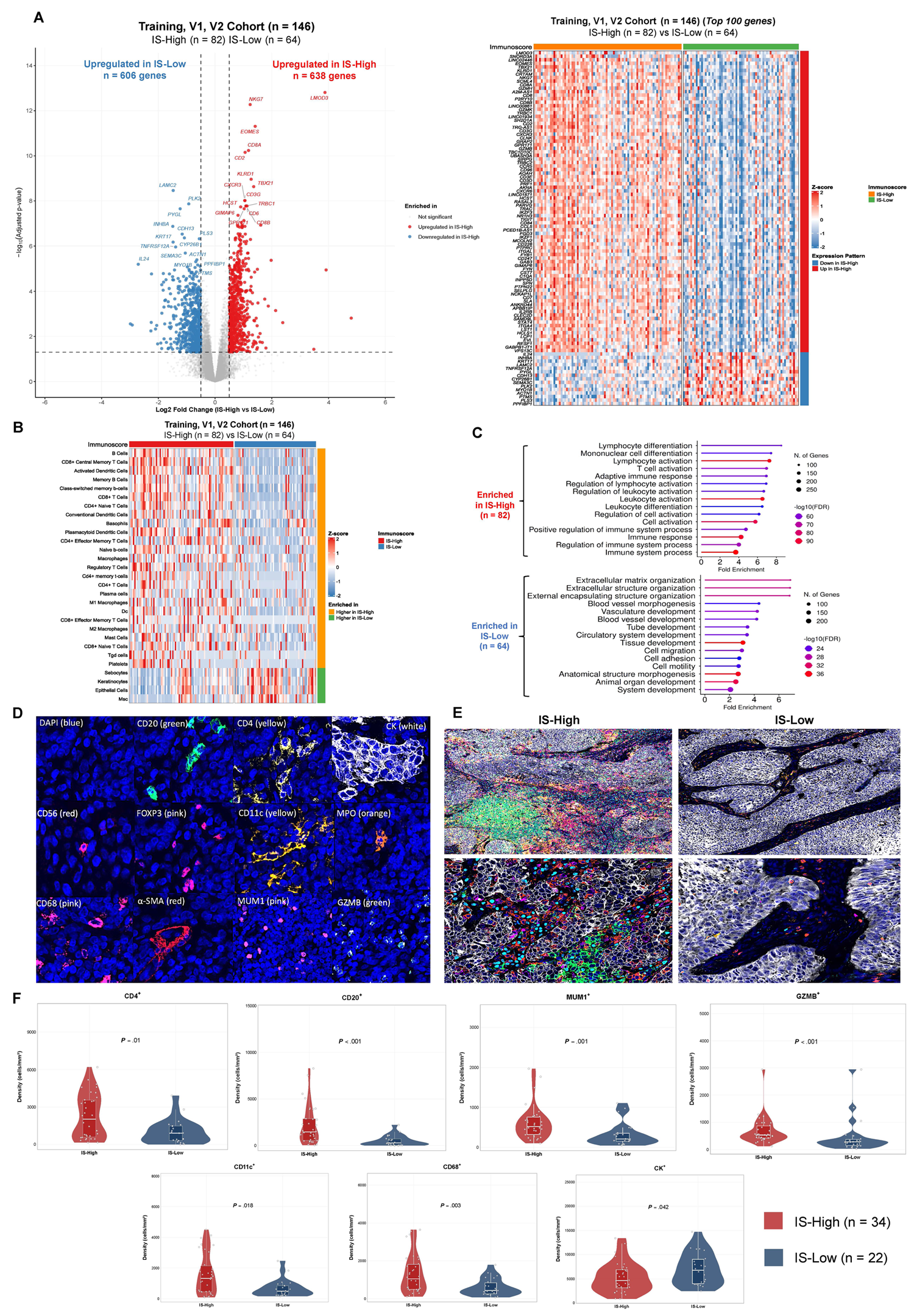
Transcriptomic and sequential immunofluorescence (seqIF) characterization of the TME in IS-High versus IS-Low patients. (A) Volcano plot and heatmap of differentially expressed genes in the Training, V1 and V2 cohorts. (B) Cell-type enrichment analysis in the Training, V1 and V2 cohorts. Dc: Dendritic cells, Tgd: gamma delta T cells, MSc: Mesenchymal stem cells. (C) GO and pathway enrichment analyses of differentially expressed genes in the Training, V1 and V2 cohorts. (D) SeqIF was performed for DAPI (cell nuclei), CD20 (B cells), CD4 (T helper cells), MUM1 (plasma cells), CD56 (NK cells), GZMB (cytotoxic cells), FOXP3 (regulatory T cells), CD11c (dendritic cells), CD68 (macrophages), cytokeratin (CK) (tumor epithelial cells), α-SMA (fibroblasts) and MPO (granulocytes). (E) Representative TME images of IS-High and IS-Low tumors: DAPI (blue), CD20□ (green), CD4□ (red), MUM1□ (cyan), CD11c□ (yellow), CD68^+^ (pink), CK□ (white). Original magnification: ×10 and ×40. (F) Mean densities of immune cells (cells/mm²) in IS-High (*N* = 34) versus IS-Low (*N* = 22) tumors.

Consistent with transcriptomic findings, seqIF confirmed a markedly inflamed TME in IS-High tumors (*N* = 34), with significantly higher densities of B cells, cytotoxic GZMB□ cells, CD4□ T cells, plasma cells, dendritic cells and macrophages (all *P* < 0.05). In contrast, IS-Low tumors (*N* = 22) showed higher densities of CK□ tumor cells (*P* = 0.042). No significant differences were observed for MPO□ granulocytes (*P* = 0.74) or α-SMA□ (myo)fibroblasts (*P* = 0.18) (**Figure 3D-F**; **Supplementary File S11**).

## DISCUSSION

In this study, we present the first multicenter, international validation of the consensus IS in HPV-associated OPSCC. Our findings demonstrate that IS is a robust prognostic biomarker for DFS, TTR and OS that significantly outperforms other clinical variables. Among 191 patients across four cohorts, IS-High patients have favorable outcomes with minimal risk of recurrence or death, whereas IS-Low patients face a poor prognosis despite standard multimodal treatment.

The prognostic value of TILs in HPV-associated OPSCC has been documented and our results are consistent with prior studies.^18,19,32–34^ Ward et al.^18^ estimated TILs based on H&E slides, stratifying patients into high- and low-risk groups with 3-year disease-specific survival rates of 96% and 59% (*P* < 0.001). Consistent with our findings, they observed that lymphocyte density was not correlated with N-stage but rather with T stage.^18^ However, their reliance on manual estimation of TILs proportions may introduce subjectivity and a risk of inter-observer variability.^21,35^ Similarly, Corredor et al.^19^ applied machine learning to digital images, finding that low levels of TILs were associated with poor DFS in Stage I HPV-related OPSCC (HR 2.27; *P* = 0.003). Similar to our results, they showed that immune infiltration, smoking status and T stage, rather than N stage, are the primary drivers of survival outcomes.^18,19^ Although informative, their approach relied solely on p16INK4a for HPV determination and the complexity of the computational workflow may hinder routine clinical implementation.^19^ By contrast, the standardized IS overcomes these limitations. Digital IS has demonstrated superior concordance and reproducibility compared with manual TILs assessment on H&E slides.^21,35^ In a comparative analysis of 540 images, Boquet et al. reported that digital software outperformed evaluations by experienced pathologists.^35^ Additionally, IS yields highly consistent results across different tissue blocks and sections from the same tumor, highlighting its analytical robustness.^20^ Importantly, relying on conventional chromogenic IHC using only two antibodies, IS is therefore cost-effective, rapid and readily implementable in most routine pathology laboratories, supporting its feasibility for widespread clinical application.^21^

The transcriptomic and seqIF data further provide strong biological validation for the IS. High CD3□/CD8□ densities detected by IHC correlated with upregulation of genes associated with a coordinated, functional cytotoxic T-cell response. Moreover, Cohort V3, analyzed using a different transcriptomic method, yielded concordant results, further supporting the robustness of our findings. (**Figure 3A–C, Supplementary Figure S9, Supplementary File S10**). Both transcriptomic and seqIF analyses consistently showed IS-High tumors enriched in CD4□ T cells, B cells, dendritic cells, and macrophages (all *P* < 0.05) (**Figure 3**). The higher B-cell signature observed may be explained by the presence of follicular helper T (Tfh) cells, which secrete CXCL13 and establish a positive loop that increases the densities of B cells, Tfh, T helper 1 and memory T cells.^36^ Concurrent enrichment of B and plasma cells also suggests potential antitumor activity via antigen presentation and antibody production. Supporting this, recent studies reported HPV-specific antibody-secreting cells in the TME of HPV-positive cancers and higher B-cell densities were associated with improved OS and enhanced responses to RT and ICIs.^34,37–39^ Similarly, the co-enrichment of dendritic cells with T and B cells in IS-High tumors also suggests the presence or early formation of tertiary lymphoid structures (TLS), which are known to support local antigen presentation and effective anti-tumor immunity.^40^ In addition, IS-High tumors were enriched in macrophages, which Ljokjel et al. reported to correlate with better 5-year disease-specific survival in OPSCC patients.^41^ Overall, our results indicate that IS not only identifies a T cell-rich TME but also an “immune-hot” IS-High phenotype, characterized by an inflamed immune landscape and associated with favorable survival outcomes.^18,19,33,34,39,42^ This highlights the practical utility of IS, which provides a reproducible measure of the complex TME without requiring costly, technically demanding multiplex IHC panels or complex transcriptomic scoring.^33,43^ Conversely, IS-Low tumors were enriched in epithelial cancer cells, mesenchymal stroma elements and angiogenesis (**Figure 3**), reflecting an immune-excluded TME that limits immune infiltration and may drive therapy resistance.^44^ Mesenchymal stromal cells can further promote tumor progression by enhancing cancer cell proliferation, angiogenesis, motility, invasion and metastasis.^45,46^

These findings directly address the ongoing de-escalation dilemma in HPV-associated OPSCC. The largely negative results of prior trials likely reflect disease heterogeneity and the limitations of current TNM- and smoking-based risk stratification.^6–12^ Although HPV-associated OPSCC is generally considered a “good-prognosis” cancer, 47.6% of patients exhibited a low IS, with a 3-year DFS of 58.9% (95% CI: 49.3-70.3) (**Figure 2B**). Transcriptomic and seqIF analyses confirmed that this subgroup represents an immune-cold TME associated with unfavorable prognosis.^44–47^ We then hypothesize that these “hidden high-risk” IS-Low patients may explain the poor outcomes seen with de-escalated treatment in prior trials.^6–12^ Given the superior performance of the combined IS + clinical model over the clinical model alone (*P* < 0.0001) (**Figure 2D**), we believe that the integration of IS could improve patient selection for future de-escalation trials. Supporting this, IS strongly stratified outcomes in both definitive RT ± CT and surgery ± adjuvant therapy settings (all *P* < 0.001), the two main contexts targeted for current dose reduction trials (**Supplementary Figure S8**).^6–12,48–50^ IS-High patients who also meet low clinical risk criteria (T1-3, N0-2 and non-active smokers; *N* = 77) may be the best candidates, given their favorable DFS, TTR and OS (**Figure 2A-B; Supplementary Figure S6-7)**. IS-High patients with high clinical risk could also be considered, as their survival was comparable to that of IS-High patients with low clinical risk; however, the limited sample size (*N* = 23) warrants validation in larger cohorts **(Supplementary Figure S7C)**. Conversely, IS-Low patients should be excluded from de-escalation or approached with more caution. Instead, they might represent a target population for treatment intensification or enhanced surveillance. Given their “cold” TME, these patients may benefit from novel strategies aimed at stimulating immune activity and converting tumors to an immune-hot phenotype, such as induction CT, ionizing RT, DNA repair-targeted therapy or vaccine-based approaches.^51^ Prospective validation of this IS-informed approach (**Supplementary Figure S12)** will be required for HPV-associated OPSCC in future clinical trials. We expect that IS will demonstrate similar utility to that observed in rectal cancer, where it successfully identified patients eligible for treatment de-escalation under the watch-and-wait strategy.^52,53^

In parallel, several studies have investigated other biomarker-driven strategies for treatment de-escalation in HPV-associated OPSCC. For instance, tumor hypoxia within the TME has been shown to impair CRT efficacy by limiting radiation-induced DNA damage and is consequently associated with poor outcomes.^54,55^ Applying this principle, Lee et al. conducted a phase II study in which tumor hypoxia was assessed using ^18^F-fluoromisonidazole (FMISO) positron emission tomography (PET) to guide de-intensified CRT in patients without evidence of hypoxia.^48^ While promising, the implementation of FMISO PET in multicenter trials remains challenging, particularly in low-resource settings, limiting its broader applicability in clinical practice. An alternative strategy was evaluated in a phase II trial by Hanna et al., which used real-time measurements of tumor tissue-modified viral (TTMV) HPV DNA to guide the selection of standard versus de-escalated CRT.^49^ Although this approach has yielded encouraging outcomes, unlike the IS, it does not allow pretreatment stratification of patient risk. Consistent with our observations, these studies highlight the critical role of biomarkers in guiding therapeutic decisions for future de-escalation trials.

Our study is subject to certain limitations. First, three of the four cohorts (Training, V1, and V3) were retrospective in design; however, validation in the prospective, multicenter V2 cohort supports the reliability of our findings. Second, although the IS cut-off was derived from the training cohort, it was based on a predefined distribution-based threshold (25^th^ percentile) rather than outcome-driven optimization, thereby limiting the risk of overfitting. The consistent performance of IS in stratifying survival across all cohorts, along with its role as the strongest independent prognostic factor in multivariable analyses, supports the robustness of the findings. Assessment of predictive accuracy using 1,000 bootstrap resamples for internal validation further confirmed the stability of IS. Although clinical data and treatment heterogeneity across cohorts reflects real-world practice, IS remained consistent, suggesting it is not overfitted to a specific dataset and has high external validity. Nevertheless, large-scale prospective validation remains warranted and is currently planned within the ongoing international, phase III de-escalation trial PATHOS (NCT02215265).^50^ IS-High patients receiving de-escalated therapy are expected to sustain favorable outcomes, supported by their strong anti-tumor immune responses, with implications for future guideline adaptation.

## CONCLUSION

Immunoscore is a robust, biologically validated prognostic tool for HPV-associated OPSCC. It effectively discriminates between patients with an excellent prognosis and those at high risk of recurrence, demonstrating superiority over current clinical staging systems. We therefore propose integrating the Immunoscore into the design of future clinical trials to evaluate its utility in guiding de-escalation strategies.

## Data Availability

All data produced in the present study are available upon reasonable request to the authors

## CONTRIBUTORS

DHN, HM and JG contributed to the conceptualization of the study and methodology. VH, NC, ASB, HKA and AH collected data and samples. DHN, MF, AM, HKA and TF performed the experiments and DHN, AM and AK carried out the analyses. DHN, HM, JG, AM and PB wrote the original draft and were responsible for validation and editing of the manuscript. All authors contributed to manuscript revision, had full access to all the data reported in the study and had final responsibility for the decision to submit the manuscript for publication.

## DATA AVAILABILITY STATEMENT

Data generated by the authors are available from the corresponding author upon reasonable request, for data types for which no community-recognized, structured repository exists.

## ACKNOWLEDGMENTS

We would like to thank the patients and their families for their participation. We also thank Jerou Aymeric and Thibaut Téo for their assistance with sample preparation.

## FIGURE LEGENDS

**Supplementary Figure S3.** Automated detection of positive cells (MUM1L plasma cells, shown in pink) was performed using HALO^TM^ software.

**Supplementary Figure S6**. Prognostic value of IS for TTR and OS. (A) Kaplan–Meier curves for TTR according to IS (High vs Low) in the overall population. (B) Kaplan–Meier curves for TTR according to the three-category IS classification (High, Intermediate, Low). (C) Kaplan–Meier curves for OS according to IS (High vs Low) in the overall population. (D) Kaplan–Meier curves for OS according to the three-category IS classification (High, Intermediate, Low).

**Supplementary Figure S7.** Prognostic value of IS across clinical risk groups. (A) Kaplan-Meier curves for DFS, TTR, and OS according to IS (High vs Low) in low-risk patients (T1-3 N0-2 non-active smokers). (B) Kaplan-Meier curves for DFS, TTR, and OS according to IS (High vs Low) in high-risk patients. (C) Kaplan–Meier curves for DFS, TTR, and OS according to combined IS and clinical risk groups. Pairwise comparisons: * *P* < 0.05; ** *P* < 0.01; *** *P* < 0.001; **** *P* < 0.0001; ns not significant.

**Supplementary Figure S8.** Prognostic value of IS for DFS, TTR, and OS across treatment subgroups: (A) Surgery + RT ± CT (*N* = 123), (B) RT ± CT (*N* = 51), and (C) Surgery only (*N* = 17).

**Supplementary Figure S9.** Volcano plot of differentially expressed genes in the V3 cohort.

**Supplementary Figure S12.** Schema of Immunoscore-informed patient stratification for future de-escalation trials in HPV-associated OPSCC.

