## Supplementary figures and images for "Validation of Immunoscore for Prognostic Stratification in HPV-associated Oropharyngeal Cancer: An International Multicenter Study"

### Supplementary Figure S6

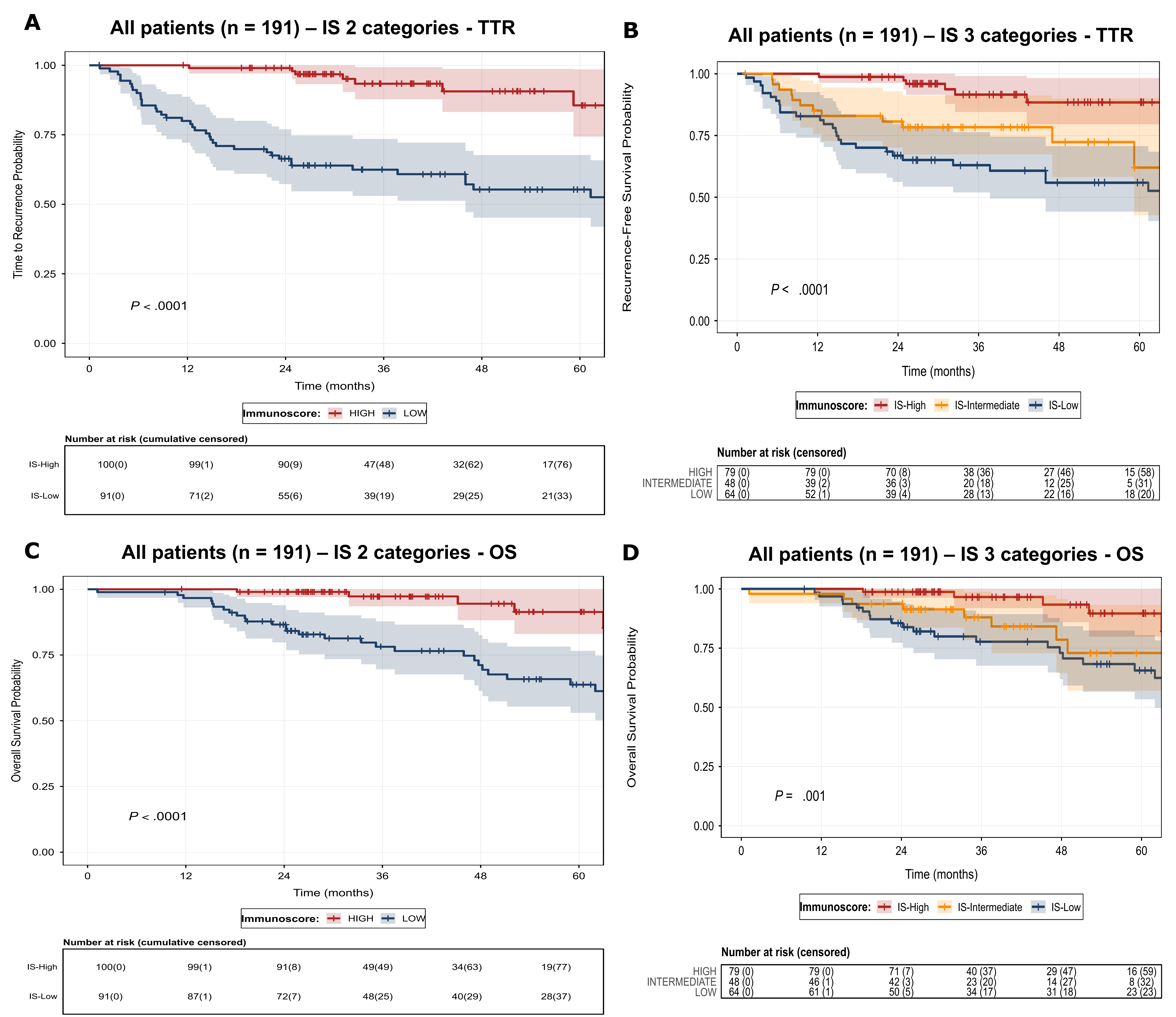

### Supplementary Figure S7

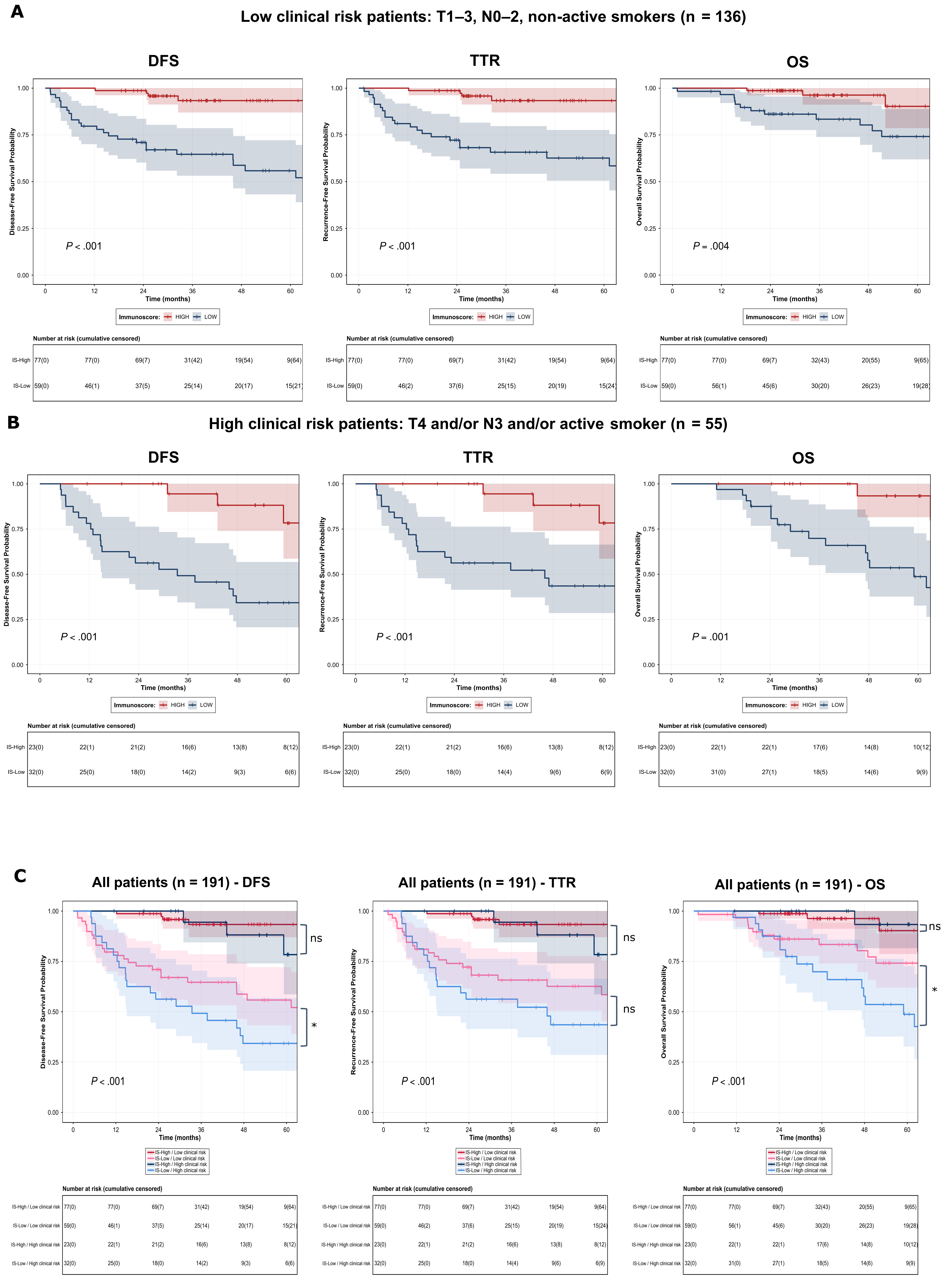

### Supplementary Figure S8

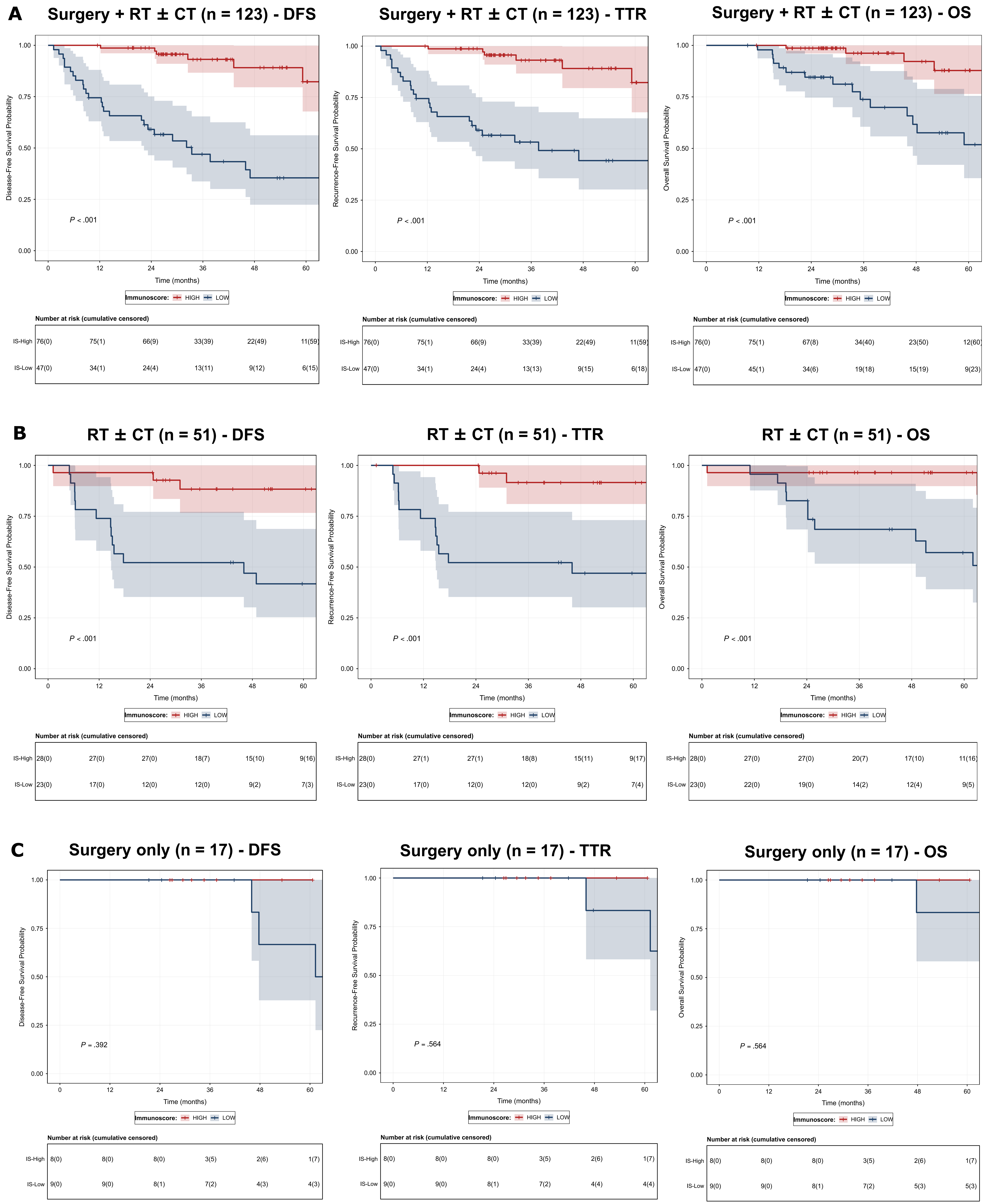

### Supplementary Figure S12

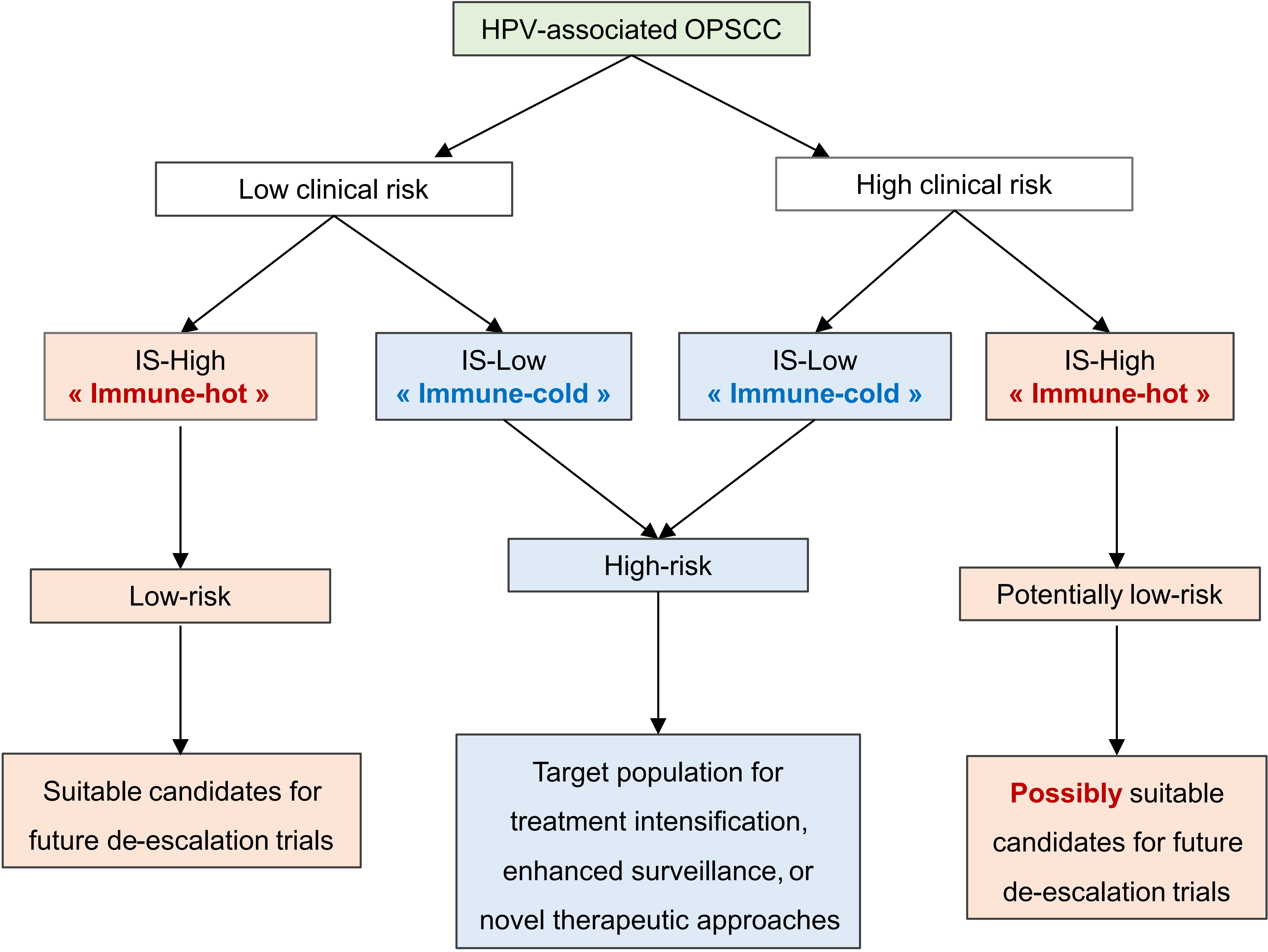
