## Supplementary File S2 for "Validation of Immunoscore for Prognostic Stratification in HPV-associated Oropharyngeal Cancer: An International Multicenter Study"

| **Marker / Cell type for IS** | **Antibody clone and manufacturer** |
| --- | --- |
| CD3+ T cells | CONFIRM anti-CD3 (2GV6) Rabbit Monoclonal Primary Antibody |
| CD8+ T cells | CD8, Monoclonal mouse antibody, C8/144B (Dako/Agilent) |

| **Marker / Cell type for seqIF** | **Antibody clone and manufacturer** |
| --- | --- |
| CD4⁺ T cells | CD4, Rabbit monoclonal, MR10020 (Lunaphore) |
| Granzyme B (GZMB)⁺ cytotoxic cells | Granzyme B, Rabbit monoclonal, EPR20129-217 (Abcam) |
| CD56⁺ natural killer cells | CD56, Rabbit monoclonal, MR10060 (Lunaphore) |
| FOXP3⁺ regulatory T cells | FOXP3, Mouse monoclonal, MR10040 (Lunaphore) |
| CD20⁺ B cells | CD20cy, Mouse monoclonal, L26 (Dako/Agilent) |
| MUM1⁺ plasma cells | MUM1p, Mouse monoclonal, M7259 (Agilent) |
| CD11c⁺ dendritic cells | CD11c, Rabbit monoclonal, MR10070 (Lunaphore) |
| CD68⁺ macrophages | CD68, Mouse monoclonal, MR10080 (Lunaphore) |
| MPO⁺ granulocytes | MPO, Polyclonal, GA511 (Dako/Agilent) |
| Pan-cytokeratin (CK)⁺ epithelial cells | Cytokeratin, Mouse monoclonal, MR10140 (Lunaphore) |
| α-SMA⁺ | α-SMA, Mouse monoclonal, MR10100 (Lunaphore) |
