## Supplementary Table S4 for "Validation of Immunoscore for Prognostic Stratification in HPV-associated Oropharyngeal Cancer: An International Multicenter Study"

| **Clinical Characteristic** | **Overall** n = 191 | **IS-High** n = 100 | **IS-Low** n = 91 | ***P*** |
| --- | --- | --- | --- | --- |
| **Age at diagnosis^*^** | 64.7 (11.0) | 63.8 (11.0) | 65.6 (11.0) | .266 |
| **Sex** |  |  |  | > .999 |
| Female | 38 (20%) | 20 (20%) | 18 (20%) |  |
| Male | 153 (80%) | 80 (80%) | 73 (80%) |  |
| **Smoking status** |  |  |  | .089 |
| Active smoker | 31 (16%) | 11 (11%) | 20 (22%) |  |
| Former smoker | 111 (58%) | 64 (64%) | 47 (52%) |  |
| Never smoker | 49 (26%) | 25 (25%) | 24 (26%) |  |
| **Smoking pack-years^+^** |  |  |  | .370 |
| ≥30 | 48 (26%) | 23 (23%) | 25 (29%) |  |
| 0-<10 | 72 (39%) | 35 (36%) | 37 (43%) |  |
| 10-<20 | 36 (19%) | 23 (23%) | 13 (15%) |  |
| 20-<30 | 29 (16%) | 17 (17%) | 12 (14%) |  |
| **T stage** |  |  |  | .12 |
| T1-2 | 126 (66%) | 71 (71%) | 55 (60.4%) |  |
| T3-4 | 65 (34%) | 29 (29%) | 36 (39.6%) |  |
| **N stage (AJCC 8^th^)** |  |  |  | .2 |
| N0 | 29 (15.2%) | 12 (12%) | 17 (18.7%) |  |
| N1-3 | 162 (84.8%) | 88 (88%) | 74 (81.3%) |  |
| **Tumor sublocalisation** |  |  |  | .561 |
| Base of Tongue | 87 (46%) | 48 (48%) | 39 (43%) |  |
| Tonsil | 104 (54%) | 52 (52%) | 52 (57%) |  |
| **Primary treatment** |  |  |  | .054 |
| RT ± CT | 51 (27%) | 23 (23%) | 28 (31%) |  |
| Surgery + CRT | 28 (15%) | 19 (19%) | 9 (10%) |  |
| Surgery + RT | 95 (50%) | 53 (53%) | 42 (46%) |  |
| Surgery only | 17 (9%) | 5 (5%) | 12 (13%) |  |
| **Follow-up time (months)**^‡^ | 37.6 (26.4- 59.7) | 35.6 (27.0- 52.7) | 41.8 (24.4- 65.6) | .478 |
| ^*^ Mean (SD)  ^+^ Data were available for 185 of 191 patients (96.9%) | | | | |

^‡^ Median (IQR, 25^th^–75^th^ percentile)
